# COVID-19, City Lockdowns, and Air Pollution: Evidence from China

**DOI:** 10.1101/2020.03.29.20046649

**Authors:** Guojun He, Yuhang Pan, Takanao Tanaka

## Abstract

The rapid spread of COVID-19 is a global public health challenge. To prevent the escalation of its transmission, China locked down one-third of its cities and strictly restricted personal mobility and economic activities. Using timely and comprehensive air quality data in China, we show that these counter-COVID-19 measures led to a remarkable improvement in air quality. Within weeks, the Air Quality Index and PM_2.5_ concentrations were brought down by 25%. The effects are larger in colder, richer, and more industrialized cities. We estimate that such improvement would avert 24,000 to 36,000 premature deaths from air pollution on a monthly basis.

## 1. Introduction

The exponential spread of the COVID-19 pandemic is a global public health crisis. In December 2019, an unknown disease, later named COVID-19, was identified in Wuhan, China (Lu et al., 2020; Zhu et al., 2020). Within three months, the disease had affected more than 100 countries (WHO, 2020). The explosion of COVID-19 cases around the world has made it a global pandemic and brought about devastating consequences (Wang et al., 2020). To contain the virus, many countries have adopted dramatic measures to reduce human interaction, including enforcing strict quarantines, prohibiting large-scale private and public gatherings, restricting private and public transportation, encouraging social distancing, imposing a curfew, and even locking down entire cities.

While the costs of enforcing these preventive measures are undoubtedly enormous, these measures could unintentionally bring about substantial social benefits. Among them, locking down cities could significantly improve environmental quality, which would partially offset the costs of these counter-COVID-19 measures. For example, satellite images caught a sharp drop in air pollution in several countries that have taken aggressive measures on the transmission of the virus.^2^

In this study, we carry out a rigorous investigation into this issue and estimate how locking down cities affected air quality at a national scale in China. We focus on China for two reasons. First, China was hit hard by the COVID-19 outbreak, and the Chinese government launched draconian countermeasures to prevent the escalation of infections (Chen et al., 2020; Kucharski et al., 2020). Nearly one-third of Chinese cities were locked down in a top-down manner, and various types of economic activities were strictly prohibited. In these cities, individuals were required to stay at home; unnecessary commercial operations and private and public gatherings were suspended; all forms of transportation were largely banned (both within a city and across cities); and mandatory temperature checking could be found in most public facilities. Second, China also suffers greatly from severe air pollution, with some estimates suggesting that air pollution is associated with an annual loss of nearly 25 million healthy life years (Kassebaum et al., 2014). If locking down cities significantly improved the air quality in China, the implied health benefits would be an order of magnitude larger than in countries with lower initial pollution levels.

Our empirical analysis uses comprehensive data at a week-by-city level from January 1^st^ to March 1^st^ in 2020. We first collect air quality data from 1,600 monitoring stations covering all the prefectural cities in China and aggregate the station level data to the city level data. We then collect the local government’s lockdown policies city by city from news media and government announcements (Appendix Table A1). Because the disease prevalence varied greatly across different regions, the terms and requirements of the lockdown also differed across provinces and cities. Thus, we define a city as locked down when all three of the following preventive measures were enforced: 1) prohibition of unnecessary commercial activities in people’s daily lives, 2) prohibition of any types of gathering by residents, 3) restrictions on private (vehicle) and public transportation. Following our definition, 95 out of 324 cities were locked down, as described in Figure 1 (Summary Statistics is described in Appendix Table A2).

**Figure 1.**
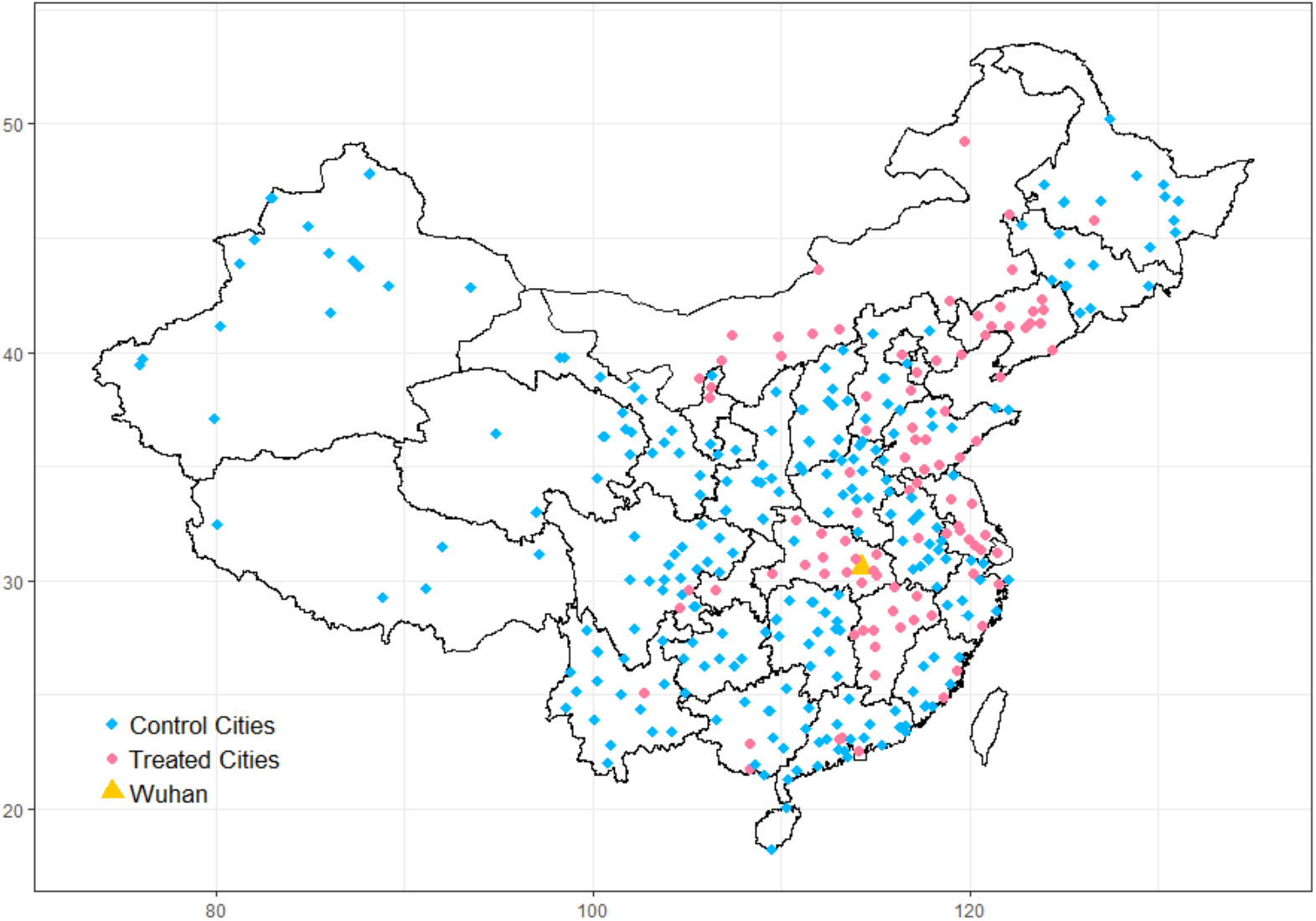
Map of the Locked-down Cities. *Notes:* This map shows which cities were locked down during the COVID-19 pandemic. The blue diamond represents locked-down cities. Overall, 95 out of 324 cities were locked down. The orange triangle indicates Wuhan city, where COVID-19 was first identified in China.

We employ two sets of difference-in-differences (DiD) models to quantify the impact of a city’s lockdown on air pollution in different groups of cities. The DiD model is a well-established econometric technique and has been widely applied in understanding the impacts of government policies (Angrist and Pischke, 2008). In our DiD specification, the city-level air pollution is as a function of the city-specific lock-down policy, city-specific fixed effects that capture city-level time-invariant determinants of air pollution (e.g., geographical conditions, baseline income), month by week fixed effects that capture shocks common to all cities in a given week (e.g., nationwide holiday policy, macroeconomic conditions) and weather conditions (see Appendix Methods). Adopting this method, we can estimate the difference in air pollution levels between the treatment group (locked-down cities) and the control group (non-locked-down cities) before and after city lockdowns in 2020. In addition, because cities without formal lockdown policies might also have been affected by the disease preventive measures (e.g., all cities extended the Spring Festival holiday, required social distancing, and urged people to stay at home), we also apply the DiD model to compare air pollution levels in the control (no-lockdown) cities before and after China’s Spring Festival relative to the previous year.^3^ Combining these two sets of DiD results, we can evaluate the total effects of city lockdown on air quality.

The underlying assumption for these DiD estimators is that treated and control cities had parallel trends in the outcome before the event. Intuitively, even if the results show that air quality improves in the locked-down city after its enforcement, the results may not be driven by the lockdown but systematic differences in treatment and control cities (e.g., treatment cities have an improving trend in air quality). Thus, we adopt the event study approach that allows for displacing the actual timing of lockdown to rule out these possibilities (Jacobson et al. 1993).

We further examine whether the effects of lockdowns vary across different types of cities. Because the lockdown restricts unnecessary industrial activities for people’s daily life, industrialized cities could be more substantially influenced by such treatments. Similarly, colder cities with higher demand for coal winter heating, richer cities with higher electricity consumption, or cities with more traffic volumes might have a more substantial impact when the lockdown is implemented.

Finally, a large strand of literature has investigated how air pollution affects population health. Based on these findings, we provide some back-of-the-envelope calculations on the potential benefits caused by air quality improvement during this period. To achieve this, we borrow estimates from recent studies that adopt a quasi-experimental approach in China and calculate the averted number of premature deaths. We focus on quasi-experimental studies because they are generally recognized as being more credible than those based on associational models (e.g., Graff Zivin and Neidell, 2013; Dominici et al., 2014). We then discuss the magnitude of the effect and the implications.

## 2. Results

### 2-1. The Trends in Air Quality

We start by presenting the patterns in the raw air quality data. In Panel A of Figure 2, we plot the AQI between the treatment and control cities over the study time in 2020. This figure shows the first margin: to what extent city lockdown affected air pollution between the treated and control cities. The figure shows the treatment group had worse air pollution levels (higher AQI) than the control group before the Chinese Spring Festival. However, the difference significantly decreased after more cities were locked down, suggesting that the lockdown improved air quality.

**Figure 2.**
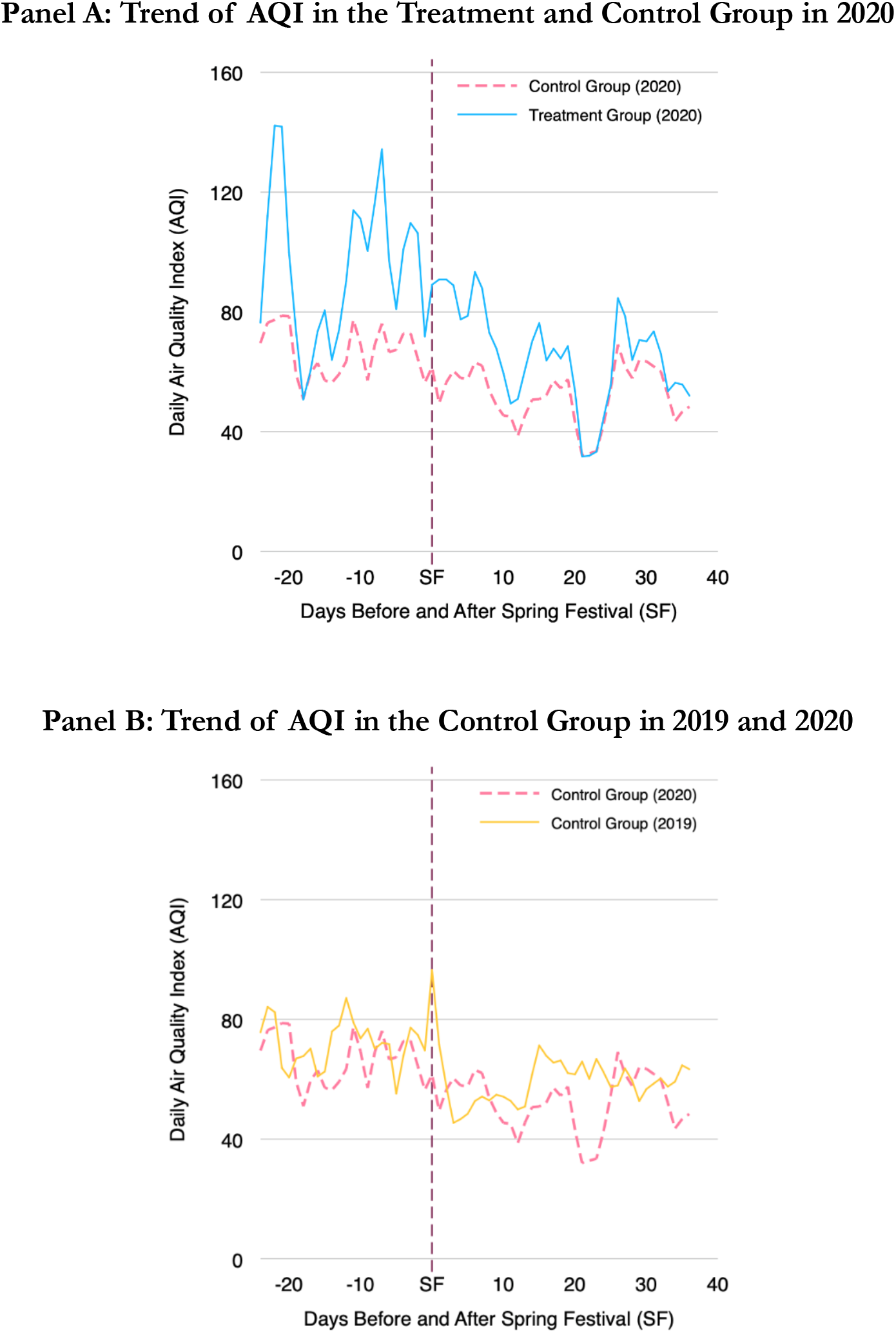
Trend of Air Quality in Treatment and Control Group. *Notes:* These figures show the trends in the Air Quality Index (AQI) in different groups of cities. In Panel A, we plot the AQI in the treatment and the control group in 2020. In Panel B, we plot the AQI in the control group in 2019 and 2020. The vertical dashed purple line represents the timing of the Chinese Spring Festival.

In Panel B of Figure 2, we look at the second margin: to what extent the control cities were also affected by the counter-virus measures. We see the AQI levels were almost equivalent before the Spring Festival in 2019 and 2020, implying the parallel trend assumption is likely to hold. In 2020, shortly after the festival, we observe that the air pollution levels became slightly lower, relative to the 2019 post-festival season. This result suggests that air quality in the control cities marginally improved, although they were not formally locked down.

### 2-2. Impacts of City Lockdown on Air Pollution

Table 1 summarizes the regression results. Here, we estimate the relative change in air pollution levels in the treatment group (locked-down cities) relative to the control group (non-locked-down cities) by fitting the DiD model described in the equation (A1). We find that a lockdown indeed improved the air quality: compared to cities without formal lockdown policies, the weekly Air Quality Index (AQI) and PM_2.5_ declined respectively by 19.4 points (18%) and 13.9 µg/m^3^ (17%) when including weather controls and a set of fixed effects (in columns (2) and (4)).^4^ These estimates are remarkably robust to the inclusion of weather variables, indicating that the changes in air pollution caused by city lockdown are not correlated with weather conditions. We also provide the results for other air pollutants (Appendix Table A3)

**Table 1.** The Effects of Lockdown on Air Quality

|  | Treatment and Control Group in 2020 |  |  |  | Control Group in 2019 and 2020 |  |  |  |
| --- | --- | --- | --- | --- | --- | --- | --- | --- |
|  | Levels |  | Log |  | Levels |  | Log |  |
|  | (1) | (2) | (3) | (4) | (5) | (6) | (7) | (8) |
| *Panel A. Air Quality Index (AQI)* |  |  |  |  |  |  |  |  |
| Lockdown | -18.84***<br>(3.27) | -19.43***<br>(3.04) | -0.17***<br>(0.03) | -0.18***<br>(0.03) |  |  |  |  |
| Spring Festival in 2020 |  |  |  |  | -6.51**<br>(2.53) | -8.77***<br>(2.40) | -0.04<br>(0.03) | -0.07***<br>(0.03) |
| R-Squared | 0.695 | 0.723 | 0.769 | 0.793 | 0.647 | 0.655 | 0.701 | 0.711 |
| *Panel B. PM<sub>2.5</sub> (μg/m<sup>3</sup>)* |  |  |  |  |  |  |  |  |
| Lockdown | -13.47***<br>(2.65) | -13.94***<br>(2.36) | -0.15***<br>(0.03) | -0.17***<br>(0.03) |  |  |  |  |
| Spring Festival in 2020 |  |  |  |  | -7.58***<br>(2.06) | -8.40***<br>(2.07) | -0.06*<br>(0.03) | -0.08**<br>(0.03) |
| R-Squared | 0.722 | 0.752 | 0.809 | 0.828 | 0.651 | 0.654 | 0.729 | 0.735 |
| Weather Control |  | Y |  | Y |  | Y |  | Y |
| City FE | Y | Y | Y | Y | Y | Y | Y | Y |
| Month-by-Week FE | Y | Y | Y | Y | Y | Y | Y | Y |
| Obs. | 2,916 | 2,916 | 2,916 | 2,916 | 4,158 | 4,158 | 4,158 | 4,158 |
| No. of Cities | 324 | 324 | 324 | 324 | 232 | 232 | 232 | 232 |

*Notes:* Weather controls include weekly mean temperature, its square, weekly mean precipitation, and weekly mean snow depth. Standard errors are clustered at the city level and reported below the coefficients. \* significant at 10% \*\* significant at 5%. \*\*\* significant at 1%.

In Columns (5) to (8), we estimate the changes in air pollution levels in the control (no-lockdown) cities before and after the Spring Festival relative to the previous year by fitting the second DiD model described in the equation (A3). We find that air quality in 2020 improved relative to the previous year’s air quality after the Festival. The results show that

AQI decreases by 8.8 points (7%) and PM_2.5_ by 8.4 (8%) after controlling for weather variables (in column ((6) and (8)), suggesting that the disease preventive measures matter for cities even without formal lockdown.

Combining these two sets of results, we estimate the total effects of city lockdown on air quality relative to the identical season in the previous year. We find that lockdown improved air quality substantially: it reduced AQI and PM_2.5_ by around 25% in the locked-down cities and 7∼8% in the control cities.

### 2-3. Tests for Pre-Treatment Prarallel Trends and Robustness Checks

We adopt the event study approach to test whether the parallel trend assumption holds, which is an assumption for DiD estimators to be valid. (Refer to Appendix Method). Figure 3 reports the regression results. (The corresponding regression results are reported in Appendix Table A4.) In Panel A, we estimate Equation (A2) and plot the estimated coefficients and their 95% confidence intervals. We find that there is no systematic difference in the trends between treatment and control groups before the city lockdown. The figure shows that the estimated coefficients for the lead terms (*k* ≤ −2) are all statistically insignificant. We also see that the trends break after the city lockdown, i.e., the lag terms (*k* ≥ 0) became negative and statistically significant. The AQI dropped by 20∼30 points within two weeks after lockdown, and this result remains statistically significant in subsequent periods.

**Figure 3.**
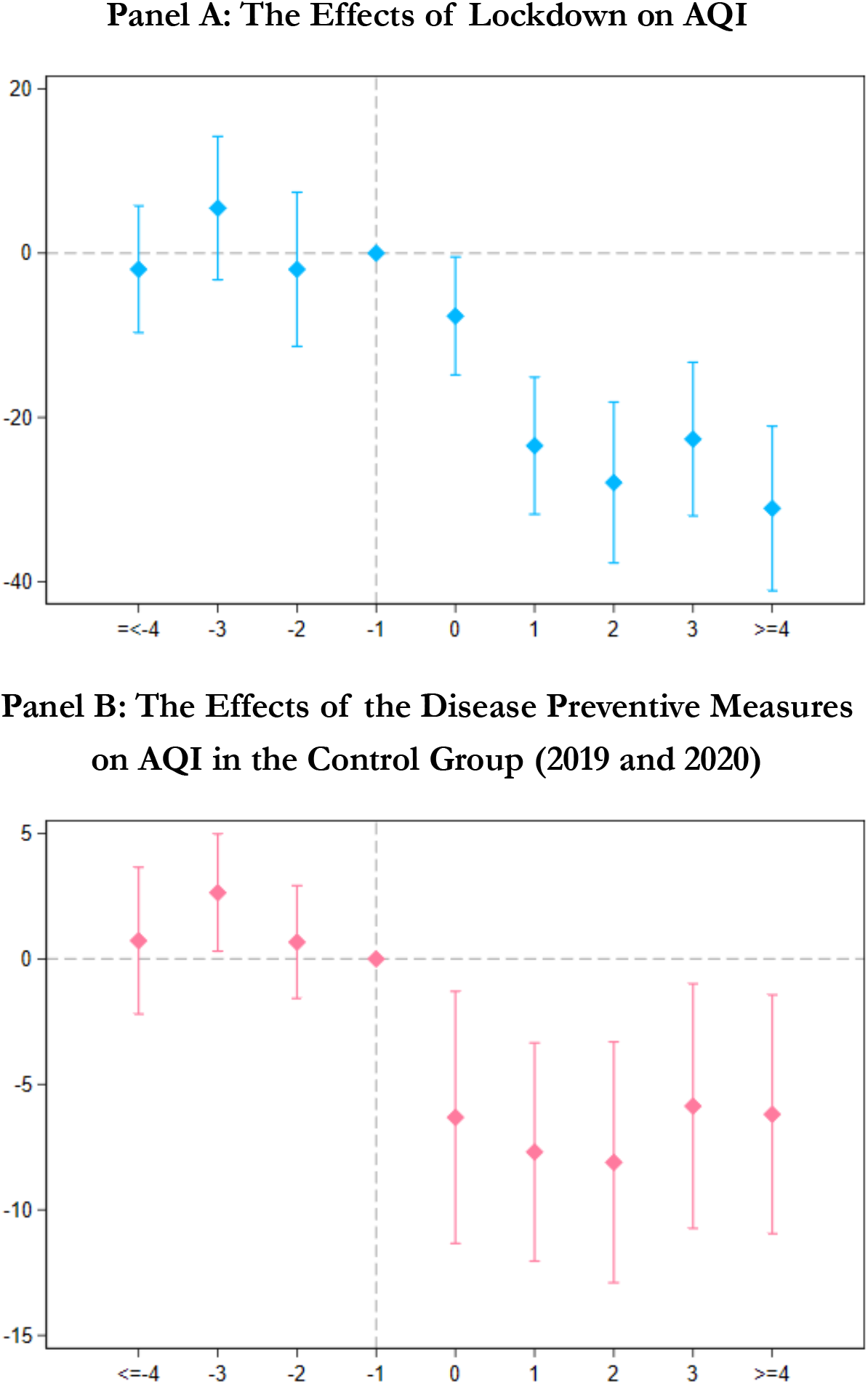
The Effects of Lockdown Before and After its Implementation. *Notes:* These figures summarize the results of the parallel trend tests. We include leads and lags of the start of the city lockdown dummy in the regressions. The dummy variable indicating one week before the city lockdown is omitted from the regressions. The estimated coefficients and their 95% confidence intervals are plotted. In Panel A, we compare the air pollution levels between the treated cities with the control cities, and in Panel B, we compare air pollution levels in the control cities between 2019 and 2020.

In Panel B, we test the parallel trend assumption for cities in the control group using data in 2019 and 2020. (The corresponding regression results are reported in Appendix Table A5.) The results suggest the air quality in 2019 could be a reasonable counterfactual for air quality in 2020 in the controlled cities; we find that their trends in air quality before the Chinese Spring Festival were also similar. The estimated coefficients after the festival show a meaningful reduction in air pollution, with the AQI being reduced by 5 to 10 points. Appendix Figure A2 repeats the same exercise using log AQI, PM_2.5,_ and log PM_2.5_ as the outcomes, and we observe very similar patterns.

Finally, we provide additional evidence that our baseline findings are not driven by specific choices we made in the empirical analyses. First, in Panel A in Appendix Table A6, we exclude cities in Hubei province, where the COVID-19 was first identified in China, and re-estimate Table 1. All the findings remain similar, suggesting that our results are not driven by a few cities that were most affected by the virus. In Panel B, we repeat this exercise using daily-level data and again reach a similar conclusion.

### 2-4. Heterogeneity

In Figure 4, we show that the effect of lockdown varies significantly across different types of cities. The impact of city lockdown on air quality was greater in colder, larger, richer, and more industrialized cities.

**Figure 4.**
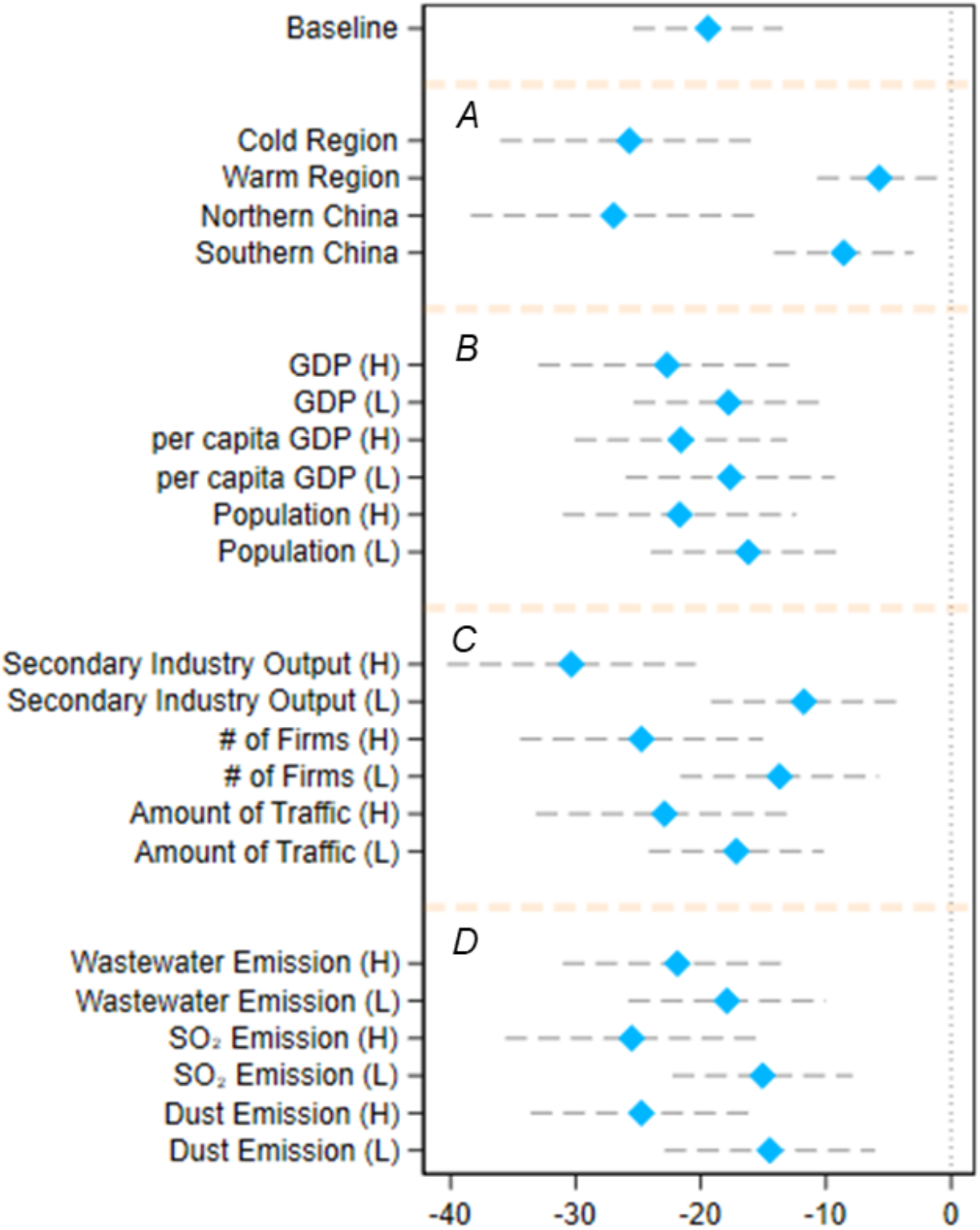
Heterogeneous Effects of Lockdown on the Air Quality Index. *Notes:* The X-axis shows the estimated coefficients and their 95% confidence intervals. Each row corresponds to a separate regression using a corresponding subsample. We use the mean values to separate the “high” group from the “low” group for each pair of heterogeneity analyses. For example, if a city’s GDP is higher than the mean GDP, it falls into a “high” GDP group. For temperature (colder or warmer group), we use data measured in the first week of our study period. North and South are divided along the Huai River. Other socio-economic data for the classification are measured in 2017. Each regression implements the equation and controls for weather, city fixed effects, and month-by-week fixed effects. Standard errors are clustered at the city level.

Panel A reveals that the impact is much more significant in colder cities or northern cities. The estimated reduction in the AQI is around 20∼30 points for those cities and is 0∼10 points in warmer or southern cities. The difference could be driven by the fact that colder cities consume more coal for heating, and lockdown reduces such consumption in offices, plants, and schools, which led to a greater improvement in air quality. In Panel B, we see the effect is greater in cities with larger GDP, population, and higher income (measured by per capita GDP). This is consistent with the fact that energy consumption is higher in more agglomerated economies, where a lot of economic activities take place. Finally, Panels C and D show that, in the cities that rely more on industrial activities (measured by the manufacturing output, the volume of traffic, the number of firms, and the emissions of different types of pollutants), the effect is more substantial.

This finding implies that coal consumption, industrial activities, and transportation all contribute substantially to air pollution in China. We repeat our heterogeneity analysis for PM_2.5_ and similarly illustrate the results in Appendix Figure A3. Appendix Table A7 presents the full set of results on AQI and PM_2.5_.

### 2-5. Potential Benefits from Improved Air Quality

A large strand of literature has investigated how air pollution affects population health. Here, we provide some back-of-the-envelope calculations on the potential benefits of the air quality improvement caused by China’s efforts to contain the virus.

The results are summarized in Table 2. Recall that the counter-COVID-19 measures reduced PM_2.5_ in the control cities by 8.40 µg/m^3^, and the city lockdown further decreased PM_2.5_ in the treated cities by 13.9 µg/m^3^(total 22.3 µg/m^3^). Two recent quasi-experimental studies in China show that a 10 µg/m^3^ increase in PM_2.5_ would lead to a 2.2%∼3.25% increase in weekly/monthly mortality (He et al., 2020; Fan et al., 2020).^5^ That implies, the total number of averted premature deaths caused by the improvement in air quality would be around 24,000 to 36,000 on a monthly basis. These numbers are significantly larger than the total number of deaths caused by COVID-19 in China (less than 3,300 as of March 28, 2020) (WHO, 2020) and illustrate the enormous social costs associated with air pollution.

**Table 2.** Estimated Health Benefits from Improved Air Quality

|  | Elasticity<br>(PM<sub>2.5</sub> per<br>10 µg/m<sup>3</sup>) | Actual<br>Change in<br>PM<sub>2.5</sub> (per<br>10 µg/m<sup>3</sup>) | Population<br>Affected<br>(per 1,000) | Base<br>Monthly<br>Mortality<br>Rate<br>(per 1000) | Estimated<br>Monthly<br>Deaths |
| --- | --- | --- | --- | --- | --- |
|  | (1) | (2) | (3) | (4) | (5) |
| *Panel A. Elasticity from Fan et al. (2020)* |  |  |  |  |  |
| Lockdown Cities | 2.20% | 2.234 | 545,820 | 0.59 | 15,939 |
| Control Cities | 2.20% | 0.84 | 738,720 | 0.59 | 8,111 |
|  |  |  |  | total | 24,050 |
| *Panel B. Elasticity from He et al. (2020)* |  |  |  |  |  |
| Lockdown Cities | 3.25% | 2.234 | 545,820 | 0.59 | 23,546 |
| Control Cities | 3.25% | 0.84 | 738,720 | 0.59 | 11,983 |
|  |  |  |  | total | 35,529 |

*Notes:* We estimate the overall monthly number of deaths in China by multiplying four variables represented in Columns (1) to (4). In Column (1), the elasticity indicates the extent to which a 10 µg/m<sup>3</sup> change in PM<sub>2.5</sub> concentration affects the mortality rate. Column (2) summarizes estimated changes in PM<sub>2.5</sub> in different types of cities based on Table 1. In Column (4), we obtain China's annual death rate from World Development Indicators and divide it by 12 to obtain the monthly death rate. As we do not have city-level mortality rate, in this calculation, we assume that the baseline mortality rate is the same across cities.

## 3. Conclusion

Using a timely and comprehensive dataset, we investigate the effect of city lockdown on air quality in China, which could bring about massive social benefits and partially offset the costs of the COVID-19 epidemic. We find that such drastic preventive measures have a significant impact on air quality. We estimated that air quality improved by around 25% (a

28.2 point decline for AQI, and 22.3µg/m^3^ for PM_2.5_) relative to the same season in 2019. We also showed that, even in the control cities, where lockdown was not fully implemented, air quality improved (AQI decreased by 8.8 points (7%) and PM_2.5_ by 8.4 µg/m^3^ (8%)). These effects are much larger in more industrialized, richer, and colder cities.

The remarkable improvement in air quality could lead to substantial health benefits. Based on the previous estimates on the relationship between air pollution and mortality, we show that averted premature deaths per month could be around 24,000 to 36,000, which is an order of magnitude larger than the number of deaths caused directly by COVID-19 in China (WHO, 2020). Because air pollution also has significant impacts on morbidity, productivity, and defensive (preventive) expenditures, our estimates should be interpreted as the lower bound of the benefits that can be derived from air quality improvement.^6^

Our findings have important policy implications for China to mitigate air pollution. We find that shutting down the unnecessary commercial activities reduces air pollution by around 25%, and the effects are even greater in cities with a larger economy, greater industrial activities and traffic, and higher demand for coal heating. These results suggest that such activities are indeed important sources of air pollution and provide a benchmark for future environmental regulation and highlight the necessity to control emissions from these sources when the business goes back to normal.

Meanwhile, although the air quality improvement during this period was unprecedented, the air pollution levels during the lockdown remained high. For example, the PM_2.5_ concentration in locked down cities was still more than four times higher than WHO considers safe (10 µg/m^3^ for the annual mean) (WHO, 2005), even though almost all non-essential production and business activities were suspended. This finding suggests that other sources of air pollution continue to contribute significantly; in particular, the coal-fired winter heating system could be the primary polluting source during our study period (Chen et al., 2013; Ebenstein et al., 2017). Our research highlights that, without further reducing its reliance on coal, it will be a real challenge for China to win its “war against pollution” (Greenstone and Schwartz, 2018; Greenstone et al., 2020).

We conclude by pointing out two caveats of this study. First, we only consider the short-term effects of city lockdown. As cities resume normal activities, the health benefits of air quality improvement could be offset in the longer term. Second, our calculation of the averted number of deaths is not based on actual mortality data, which are not yet available. If COVID-19 or city lockdown affects mortality through other channels, the overall mortality costs could be higher or lower, depending on how different channels are affected. For example, medical resources in many cities ran short immediately after the disease outbreak, thus patients could die because they were unable to receive timely and proper treatment (Xie et al., 2020). The counter-virus measures also negatively affected the economy and employment, which are detrimental to population health. In such cases, excess deaths could be caused by economic consequences than were saved by reduced pollution. In contrast, as the pandemic also signficantly increased individuals’ awareness of their health conditions and made people practice good hygiene, the counter-virus measures might reduce deaths from other diseases, particularly influenza.^7^ While estimating the overall mortality impact of COVID-19 and city lockdown is beyond the scope of our paper, future research on these issues is warranted to understand the full implications of the COVID-19 pandemic.

## Data Availability

The data that support the findings of this study are available upon reasonable request.

## Author Contributions

All authors (GH, PY, and TT) equally contributed to the paper. GH, PY, and TT conceptualized the study and carried out initial planning. PY retrieved and constructed the data set for analysis. PY constructed the summary and descriptive statistics. PY, TT, and GH designed the statistical method. PY carried out the statistical analysis, and TT conducted back-of-the-envelope calculations, which were refined by GH for the final version. TT prepared the first draft of the report, which was revised by PY and GH. All authors reviewed and contributed to a final draft and approved the final version for publication.

## Data Availability Statement

All data will be available at public repository to reproduce the results presented in this paper upon publication.

## Code Availability Statement

All replication codes will be available at public repository to reproduce the results presented in this paper upon publication.

## Online Appendix

### Appendix 1: Materials and Method

#### 1-1. Data Appendix

##### Air Quality Data

The air quality data is a high-frequency dataset covering seven major sets of air pollutants. We obtain these data from the Ministry of Ecology and Environment. The original dataset includes hourly readings on Air Quality Index (AQI), PM_2.5_, PM_10_, SO_2_, O_3_, NO_2_, and CO from 1605 air quality monitoring stations covering all the prefectural cities in China. Based on weights of the inverse of squared distance from the station and city population center, we collapse the dataset to 324 cities at a weekly level.

##### Weather Data

Weather data includes temperature, precipitation, and snow. These data are obtained from the Global Historical Climatology Network (GHCN) from the National Oceanic and Atmospheric Administration (NOAA). We collapse data to a weekly city-level dataset using the same methods as the air quality data.

##### Lockdown

We collect local governments’ lockdown information city by city from news media and government announcements. Most of the cities’ lockdown policies were directly issued by the city-level governments, while a few were promulgated by the provincial governments. To improve policy compliance, civil servants and volunteers were assigned to communities, firms, business centers, and traffic checkpoints. The local government also penalized offenders if the rules were violated. There are some variations in rules and degrees of the lockdown. For example, in some cities, individuals were not allowed to go out (food and daily necessities were delivered to them), while in other cities, they could go out if they did not have a fever. In this paper, we define lockdown when the following three measures are all enforced: 1) prohibition of unnecessary commercial activities for people’s daily lives, 2) prohibition of any type of gathering by residents, 3) restrictions on private (vehicles) and public transportation. The primary dataset for lockdown is at a daily level. Thus, we aggregate this to a weekly level. Here, we define treatment = 1 if more than half of the days in the week were locked down. The timings of the cities’ lockdowns are presented in Appendix Table A1.

##### Socio-Economic Status

To explore the heterogeneity, we assemble the cities’ socio-economic status from the 2017 China City Statistical Yearbook. It contains city-level statistics such as GDP, population, industrial output, number of firms, amount of traffic, and pollutant emissions.

##### Summary Statistics

We report the summary statistics of air pollution and weather variables during this period in Appendix Table A2. The average AQI is 74, with a standard deviation of 42. The PM_2.5_ concentration is 52 µg/m^3^, five times higher than the WHO standard (10 µg/m^3^ for annual mean, and 25 µg/m^3^ for a daily mean). Cities that were locked down were, on average, more polluted than the control cities before the lockdowns. This is likely because Wuhan and its neighboring cities are generally more polluted than cities that are far away. We also see a sharp decline in AQI after the lockdown.

### 1-2. Method

#### Difference-in-Differences Model and Event Study

We use two sets of Difference-in-Differences (DiD) models to identify the impact of counter-COVID-19 on air pollution. First, in our baseline regression, we estimate the relative change in air pollution levels between the treated and control cities using the following model:

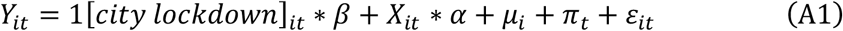

where *Y*_*it*_ represents the level of air pollution in city *i* in month-by-week *t*. 1[*city lockdown*]_*it*_ denotes whether a lockdown is enforced *i* in city in month-by-week *t*, and takes the value one if the city is locked down and zero otherwise. *X*_*it*_ are the control variables, including temperature, temperature squared, precipitation, and snow depth. *μ*_*i*_ indicate city-fixed effects and *π*_*t*_ indicate month-by-week fixed effects.

The city fixed effects, *μ*_*i*_, which are a set of city-specific dummy variables, can control for time-invariant confounders specific to each city. For example, the city’s geographical conditions, short-term industrial and economic structure, income, and natural endowment can be controlled by introducing the city fixed effects. The month-by-week fixed effects, *π*_*t*_, are a set of dummy variables that account for shocks that are common to all cities in a given week, such as the nationwide holiday policies, macroeconomic conditions, and the national air pollution time trend.

As both location and time fixed effects are included in the regression, the coefficient β estimates the difference in air pollution between the treated (locked down) cities and the control cities before and after the enforcement of the lockdown policy. We expect the coefficient β to be negative, as the industrial and business activities were restricted in the locked-down cities, and thus their air pollution levels should significantly decrease.

The underlying assumption for the DiD estimator is that treated and control cities had parallel trends in the outcome before the lockdowns. To test this assumption, we conduct a parallel trend test following Jacobson et al. (1993):

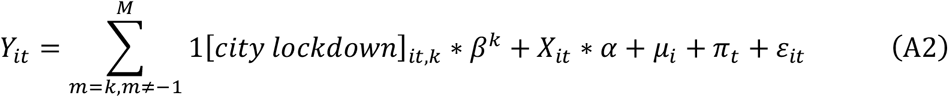

where 1[*city lockdown*]_*i,tk*_ are a set of dummy variables indicating the treatment status at different periods. The dummy for m = −1 is omitted in Equation (A2) so that the post-lockdown effects are relative to the period immediately before the launch of the policy. The parameter of interest *β*^*k*^ estimates the effect of city lockdown m weeks after the implementation. We include leads of the treatment dummy in the equation, testing whether the treatment affects the air pollution levels before the launch of the policy. Intuitively, the coefficient *β*^*k*^ measures the difference in air quality between cities under lockdown and otherwise in period *K* relative to the difference one year before the lockdown. We expect lockdown would improve air quality with *β*^*k*^ being negative when *k* ≥ 0. If the parallel trend assumption holds, *β*^*k*^ would be close to zero when *k* ≤ −2.

Even in a city that did not have a formal lockdown policy, people’s daily lives could still be affected by the counter-virus measures. In fact, in all Chinese cities, the Spring Festival holiday was extended, and people were advised to stay at home when possible, enforce social distancing, and keep good hygiene. We examine this possibility by comparing the air pollution changes between 2019 and 2020 for the same period (January 1^st^ to March 1^st^) within the control group. As the explosion of the COVID-19 cases coincided with China’s Spring Festival (SF), we investigate whether the trend of air quality in 2020 differs from the trend in 2019 after the festival, by fitting the following model:

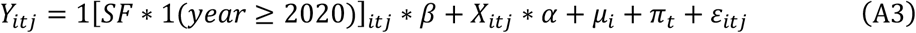

where *i* denotes city, *t* is month-by-week, and *j* represents year. 1[*SP* ∗ 1(*year* ≥ 2020)]_*it*j_ is our variable of interest, and it takes the value one if it is after the Chinese Spring Festival in the year 2020, and zero otherwise. The estimated coefficient *β* would be zero if the coronavirus and countermeasures do not affect control cities. As in Model (1), *X*_*it*j_ are the control variables, *μ*_*i*_ are city fixed effects and *π*_*t*_ indicates month-by-week fixed effects.

The identifying assumption for Model (3) is similar to Model (1). We require the trends in air quality before the Spring Festival in 2019 to be similar to the trends in air quality in the corresponding period in 2020 (i.e., the parallel trend assumption). We can test this assumption analogously using Model (2). In all the regressions, we cluster the standard errors at the city level.

#### Back-of-The-Envelope Calculations on Potential Health Benefits

We predict the reduced mortality from the improvement in air quality by calculating the following equation:

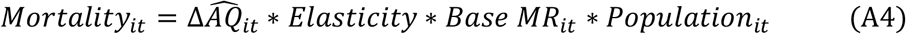

where *Mortality*_*it*_ represents estimated saved deaths in city *i* in month *t*, and 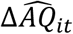 indicates an estimated change in air quality from Equations (A1) and (A3) in city *i* in month *t*, where we assume the effect of lockdown on air quality is identical within the treatment group and within the control group. We borrow the *Elasticity* estimates from existing studies that measure the effect of air quality on mortality; this represents the change in number of deaths in response to a one-unit change in air quality. *Base MR*_*it*_ represents the base mortality rate, and *Population*_*it*_ denotes the population in city *i* in week *t*. The computations are summarized in Table 2 in the main text.

For *Elasticity* estimates, we focus on studies that meet the following two criteria. First, we rely on quasi-experimental studies that investigate how exogenous changes in air quality affect health outcomes. Because the air pollution variation could be confounded by factors that also affect population health, the estimates from quasi-experimental studies are generally recognized as being more credible than those based on associational models (e.g., Graff Zivin and Neidell, 2013; Dominici et al., 2014). Second, we focus on the most recent research in China. Because the effects of air pollution on mortality could depend on the baseline pollution levels, income, and institutional quality, studies using data from earlier years may be less relevant to our research context (Arceo et al., 2014; Cheung et al., 2020).

We search for the literature and identify two studies that meet both criteria: He et al. (2020) use seasonal agricultural straw burning as the instrumental variable for PM_2.5_, and estimate how PM_2.5_ affects mortality; and Fan et al. (2020) examine how turning on the coal-fired winter heating system affects weekly mortality.^8^ These studies estimate that a 10 µg/m^3^ increase in PM_2.5_ would lead to a 2.2%∼3.25% increase in mortality, which is largely consistent with a recent quasi-experimental study in the U.S. (Deryugina et al., 2020).^9^

---

**Figure A1.**
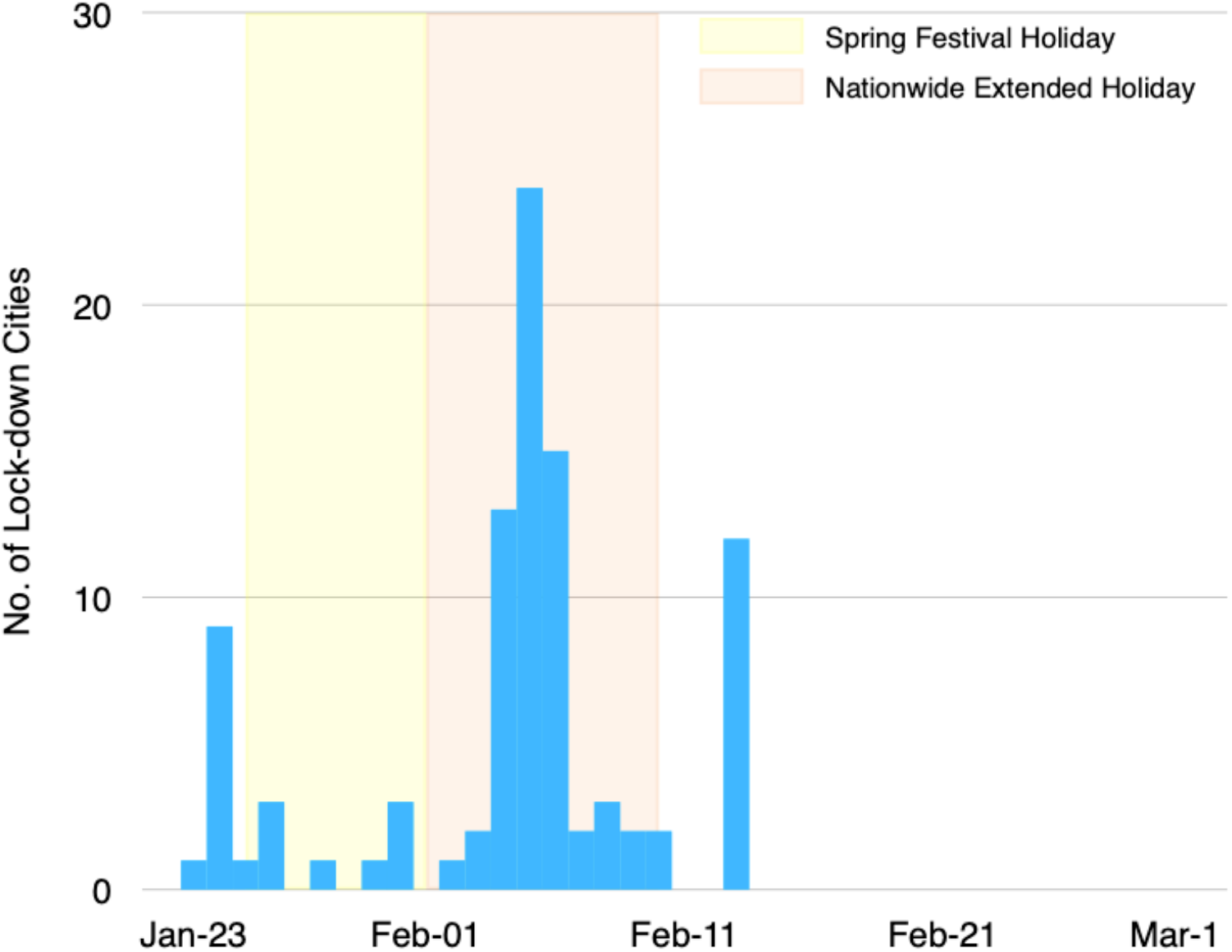
Timing of Lockdown. *Notes:* This graph shows the timing of the start of the city lockdown. The x-axis represents the date, and the y-axis represents the number of locked down cities. The yellow background represents the Chinese Spring Festival, and the red background represents the extended Spring Festival.

**Figure A2.**
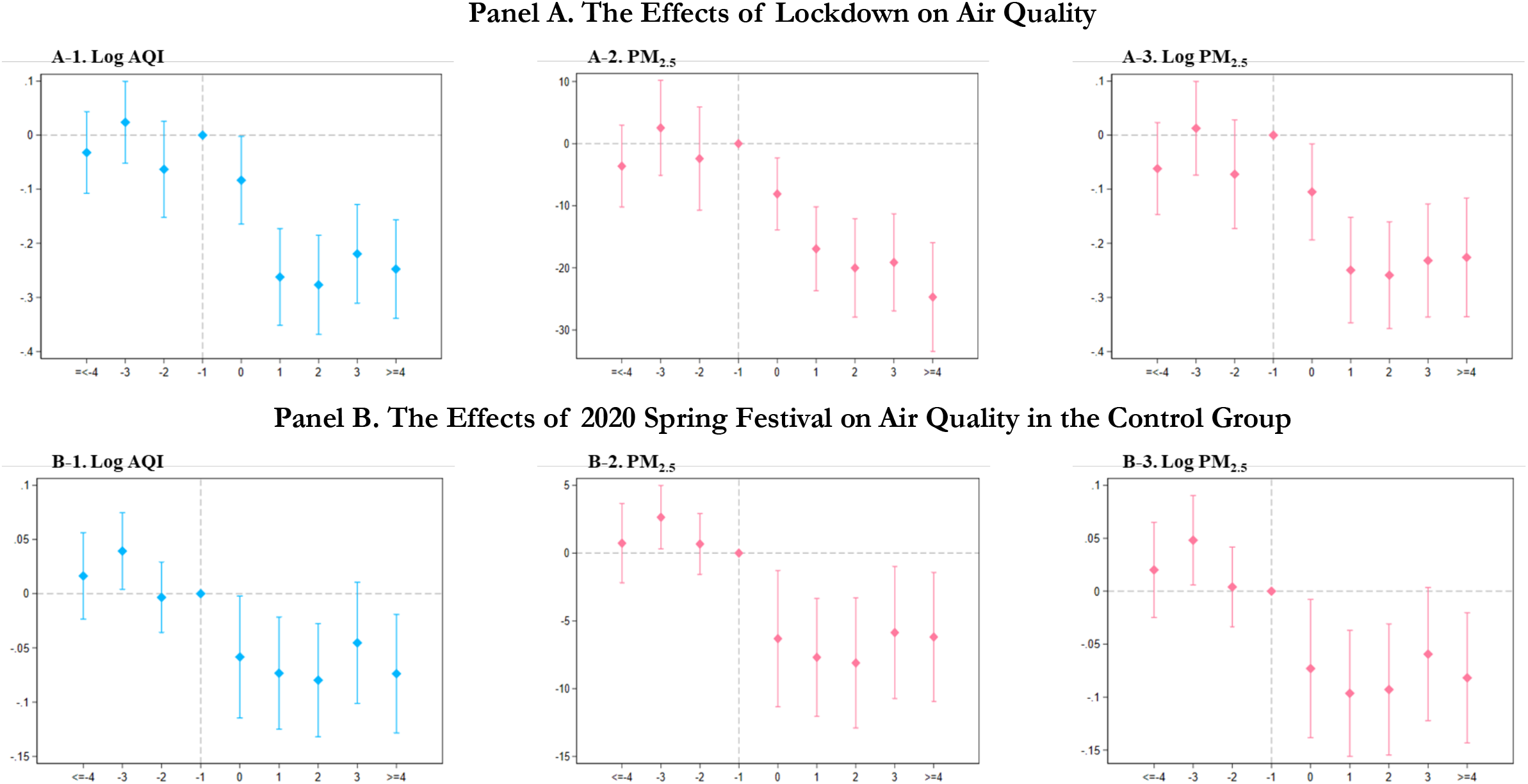
Event-Study Results on AQI and PM_2.5_ Panel A. The Effects of Lockdown on Air Quality. *Notes:* These figures summarize the results of the parallel trend tests. We include leads and lags of the start of the city lockdown dummy in the regressions. The dummy variable indicating one week before the city lockdown is omitted from the regressions. The estimated coefficients and their 95% confidence intervals are plotted. In Panel A, we compare the air pollution levels between the treated cities and the control cities, and in Panel B, we compare air pollution levels in the control cities between 2019 and 2020.

**Figure A3.**
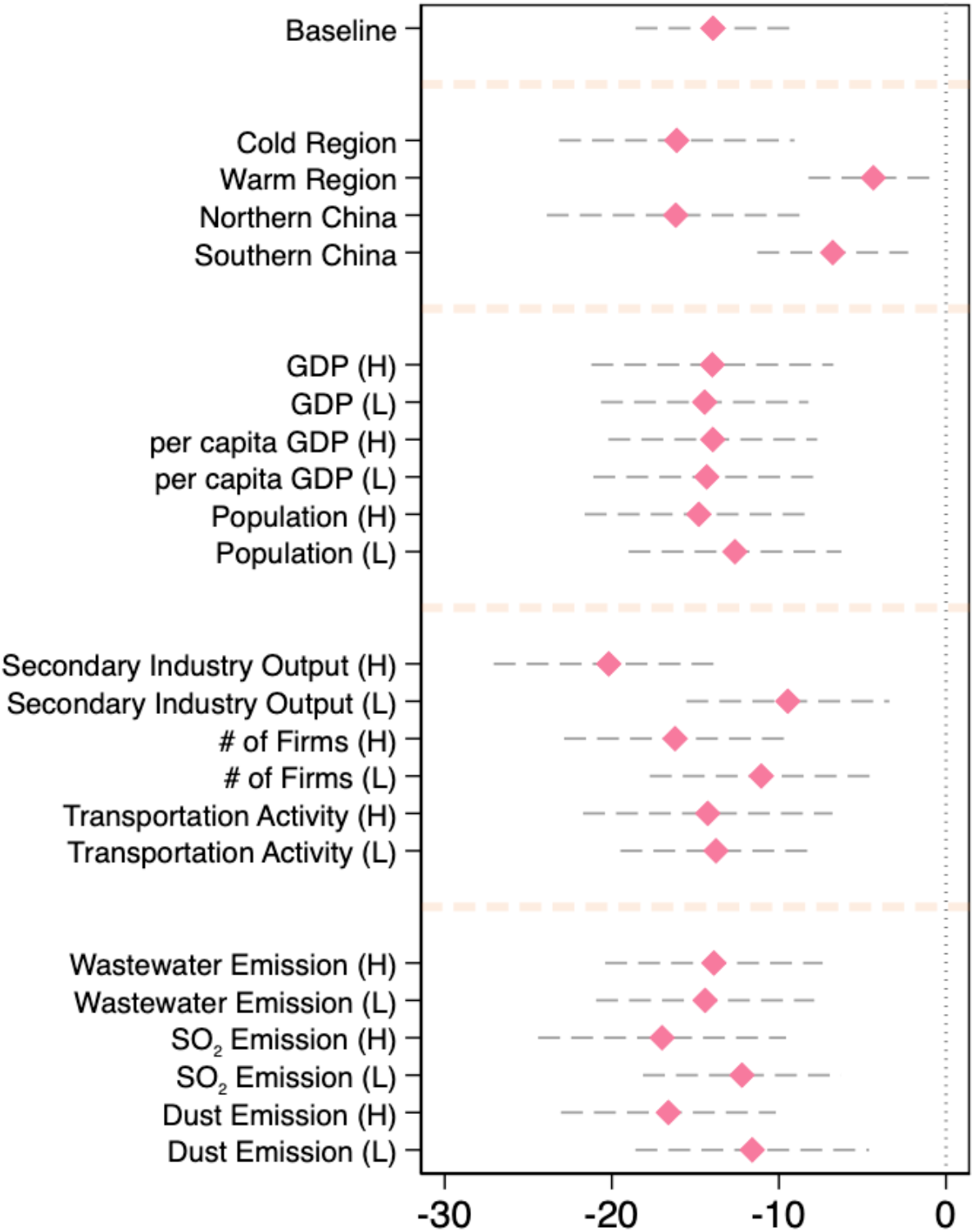
Heterogeneous Impact Using PM_2.5_. *Notes:* The x-axis shows the estimated coefficients and their 95% confidence intervals. Each row corresponds to a separate regression using a corresponding subsample. We use the mean values to separate the “high” group from the “low” group for each pair of heterogeneity analyses. For example, if a city’s GDP is higher than the mean GDP, it falls into the “high” GDP group. For temperature (colder or warmer group), we use data measured in the first week of our study period. North and South are divided along the Huai River. Other socio-economic data for the classification are measured in 2017.

**Table A1.** List of Locked Down Cities

| Starting Date | Cities |
| --- | --- |
| 23-Jan | Wuhan |
| 24-Jan | Huangshi, Shiyan, Yichang, Ezhou, Jingmen, Xiaogan, Huanggang, Xianning, Enshi |
| 25-Jan | Qinhuangdao |
| 26-Jan | Xiangyang, Jingzhou, Xiantao |
| 28-Jan | Tangshan |
| 30-Jan | Dongying |
| 31-Jan | Chongqing, Yinchuan, Wuzhong |
| 2-Feb | Wenzhou |
| 3-Feb | Wuxi, Jining |
| 4-Feb | Harbin, Nanjing, Xuzhou, Changzhou, Nantong, Hangzhou, Ningbo, Fuzhou, Jingdezhen, Zaozhuang, Linyi, Zhengzhou, Zhumadian |
| 5-Feb | Shenyang, Dalian, Anshun, Fushun, Benxi, Dandong, Jinzhou, Fuxin, Liaoyang, Panjin, Tieling, Chaoyang, Huludao, Yangzhou, Hefei, Quanzhou, Nanchang, Jinan, Qingdao, Taian, Rizhao, Laiwu, Nanning |
| 6-Feb | Tianjin, Shijiazhuang, Suzhou, Pingxiang, Jiujiang, Xinyu, Yingtan, Ganzhou, Ji'an, Yichun, Fuzhou, Shangrao, Neijiang, Yibin, Xinyang |
| 7-Feb | Suzhou, Guangzhou |
| 8-Feb | Shenzhen, Foshan, Fangchenggang, |
| 9-Feb | Cangzhou, HuaiBei |
| 10-Feb | Beijing, Shanghai |
| 13-Feb | Hohhot, Baotou, Wuhai, Chifeng, Tongliao, Ordos, Hulun Buir, Bayan Nur, Ulanqab, Xing'an League, Xilingol League, Alxa League |

*Notes:* The lockdown information is from local government and various media news in 2020.

**Table A2.**
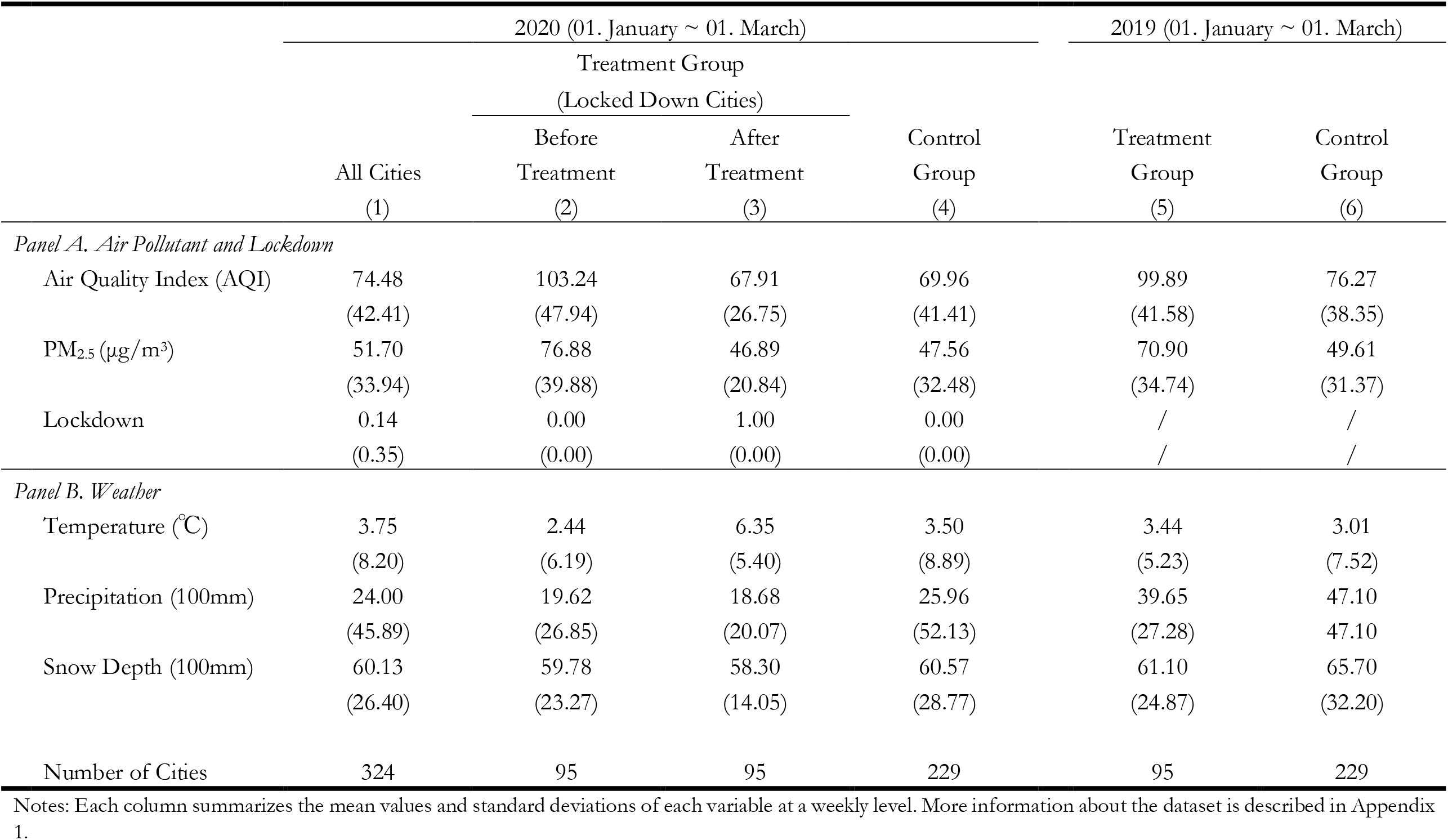
Summary Statistics

**Table A3.** Main Specification Using Other Air Pollutants

|  | Treatment and Control Cities in 2020 |  |  |  |  |
| --- | --- | --- | --- | --- | --- |
|  | CO | NO<sub>2</sub> | PM<sub>10</sub> | SO<sub>2</sub> | O<sub>3</sub> |
|  | (mg/m<sup>3</sup>) | (µg/m<sup>3</sup>) | (µg/m<sup>3</sup>) | (µg/m<sup>3</sup>) | (µg/m<sup>3</sup>) |
|  | (1) | (2) | (3) | (4) | (5) |
| *Panel A. Levels* |  |  |  |  |  |
| Lockdown | -0.07***<br>(0.03) | -4.70***<br>(0.63) | -22.22***<br>(3.18) | -1.25**<br>(0.49) | 3.10***<br>(0.76) |
| R-Squared | 0.788 | 0.870 | 0.688 | 0.857 | 0.770 |
| *Panel B. log* |  |  |  |  |  |
| Lockdown | -0.03***<br>(0.01) | -0.13***<br>(0.02) | -0.22***<br>(0.03) | -0.09***<br>(0.02) | 0.09***<br>(0.02) |
| R-Squared | 0.814 | 0.907 | 0.811 | 0.907 | 0.753 |
| Weather Control | Y | Y | Y | Y | Y |
| City FE | Y | Y | Y | Y | Y |
| Date FE | Y | Y | Y | Y | Y |
| Obs. | 2,916 | 2,916 | 2,916 | 2,916 | 2,916 |
| No. of Cities | 324 | 324 | 324 | 324 | 324 |

*Notes:* Weather controls include weekly mean temperature, its square, weekly mean precipitation, and weekly mean snow depth. Standard errors are clustered at the city level and reported below the coefficients. \* significant at 10% \*\* significant at 5%. \*\*\* significant at 1%.

**Table A4.** Event Study: The Effects of Lockdown on Air Quality

|  | AQI |  | PM<sub>2.5</sub> (μg/m<sup>3</sup>) |  |
| --- | --- | --- | --- | --- |
|  | levels<br>(1) | log<br>(2) | levels<br>(3) | log<br>(4) |
| >=4 Weeks Before | -1.97<br>(3.92) | -0.03<br>(0.04) | -3.62<br>(3.35) | -0.06<br>(0.04) |
| 3 Weeks Before | 5.48<br>(4.43) | 0.02<br>(0.04) | 2.54<br>(3.89) | 0.01<br>(0.04) |
| 2 Weeks Before | -1.97<br>(4.77) | -0.06<br>(0.05) | -2.41<br>(4.23) | -0.07<br>(0.05) |
| Treatment Week | -7.65**<br>(3.65) | -0.08**<br>(0.04) | -8.10***<br>(2.95) | -0.11**<br>(0.05) |
| 1 Week Later | -23.45***<br>(4.24) | -0.26***<br>(0.05) | -16.94***<br>(3.43) | -0.25***<br>(0.05) |
| 2 Weeks Later | -27.93***<br>(4.97) | -0.28***<br>(0.05) | -20.01***<br>(4.02) | -0.26***<br>(0.05) |
| 3 Weeks Later | -22.65***<br>(4.75) | -0.22***<br>(0.05) | -19.13***<br>(3.97) | -0.23***<br>(0.05) |
| >=4 Weeks Later | -31.09***<br>(5.09) | -0.25***<br>(0.05) | -24.70***<br>(4.45) | -0.23***<br>(0.06) |
| Weather | Y | Y | Y | Y |
| City FE | Y | Y | Y | Y |
| Month-by-Week FE | Y | Y | Y | Y |
| Obs. | 2,916 | 2,916 | 2,916 | 2,916 |
| R-Squared | 0.719 | 0.795 | 0.740 | 0.822 |
| No. of Cities | 324 | 324 | 324 | 324 |

*Notes:* Weather controls include weekly mean temperature, its square, weekly mean precipitation, and weekly mean snow depth. Standard errors are clustered at the city level and reported below the coefficients. \* significant at 10% \*\* significant at 5%. \*\*\* significant at 1%

**Table A5.** Event Study: The 2020 Spring Festival on Air Quality in The Control Group

|  | AQI |  | PM<sub>2.5</sub> (µg/m<sup>3</sup>) |  |
| --- | --- | --- | --- | --- |
|  | levels<br>(1) | log<br>(2) | levels<br>(3) | log<br>(4) |
| >=4 Weeks Before | 0.65<br>(1.95) | 0.02<br>(0.02) | 0.73<br>(1.48) | 0.02<br>(0.02) |
| 3 Weeks Before | 2.46*<br>(1.48) | 0.04**<br>(0.02) | 2.64**<br>(1.19) | 0.05**<br>(0.02) |
| 2 Weeks Before | 0.31<br>(1.51) | -0.00<br>(0.02) | 0.67<br>(1.14) | 0.00<br>(0.02) |
| Treatment Week | -6.21**<br>(2.94) | -0.06**<br>(0.03) | -6.32**<br>(2.55) | -0.07**<br>(0.03) |
| 1 Week Later | -8.18***<br>(2.67) | -0.07***<br>(0.03) | -7.70***<br>(2.21) | -0.10***<br>(0.03) |
| 2 Weeks Later | -8.65***<br>(2.78) | -0.08***<br>(0.03) | -8.10***<br>(2.44) | -0.09***<br>(0.03) |
| 3 Weeks Later | -5.70*<br>(2.96) | -0.05<br>(0.03) | -5.87**<br>(2.47) | -0.06*<br>(0.03) |
| >=4 Weeks Later | -7.96***<br>(3.01) | -0.07***<br>(0.03) | -6.19**<br>(2.42) | -0.08***<br>(0.03) |
| Weather | Y | Y | Y | Y |
| City FE | Y | Y | Y | Y |
| Month-by-Week FE | Y | Y | Y | Y |
| Obs. | 4,158 | 4,158 | 4,158 | 4,158 |
| R-Squared | 0.653 | 0.712 | 0.652 | 0.735 |
| No. of Cities | 232 | 232 | 232 | 232 |

*Notes:* Weather controls include weekly mean temperature, its square, weekly mean precipitation, and weekly mean snow depth. Standard errors are clustered at the city level and reported below the coefficients. \* significant at 10% \*\* significant at 5%. \*\*\* significant at 1%

**Table A6.** Robustness Check Using Different Samples

|  | AQI | log (AQI) | PM<sub>2.5</sub> | log (PM<sub>2.5</sub>) |
| --- | --- | --- | --- | --- |
|  | (1) | (2) | (3) | (4) |
| *Panel A. Drop Cities in Hubei Province* |  |  |  |  |
| Lockdown | -21.69***<br>(3.3) | -0.20***<br>(0.03) | -15.83***<br>(2.56) | -0.19***<br>(0.03) |
| R-Squared | 0.725 | 0.796 | 0.754 | 0.831 |
| Obs. | 2808 | 2808 | 2808 | 2808 |
| Month-by-Week FE | Y | Y | Y | Y |
| *Panel B. Using Daily Data* |  |  |  |  |
| Lockdown | -19.84***<br>(3.13) | -0.17***<br>(0.03) | -14.07***<br>(2.53) | -0.17***<br>(0.03) |
| R-Squared | 0.515 | 0.601 | 0.541 | 0.641 |
| Obs. | 19764 | 19764 | 19764 | 19764 |
| Date FE | Y | Y | Y | Y |
| Weather Control | Y | Y | Y | Y |
| City FE | Y | Y | Y | Y |
| No. of Cities | 324 | 324 | 324 | 324 |

*Notes:* Weather controls include mean temperature, its square, precipitation, and snow depth. Standard errors are clustered at the city level and reported below the coefficients. \* significant at 10% \*\* significant at 5%. \*\*\* significant at 1%.

**Table A7.** Heterogeneity Analysis

|  | AQI |  | PM<sub>2.5</sub> (μg/m<sup>3</sup>) |  |
| --- | --- | --- | --- | --- |
|  | levels | log | levels | log |
|  | (1) | (2) | (3) | (4) |
| *Panel A* |  |  |  |  |
| Cold Region | -26.97*** | -0.20*** | -16.17*** | -0.12*** |
| (Obs. = 1,224) | (5.77) | (0.05) | (3.90) | (0.04) |
| Warm Region | -8.58*** | -0.12*** | -6.78*** | -0.14*** |
| (Obs. = 1,692) | (2.83) | (0.03) | (2.29) | (0.04) |
| Northern China | -25.71*** | -0.20*** | -16.11*** | -0.13*** |
| (Obs. = 1,413) | (5.23) | (0.04) | (3.57) | (0.04) |
| Southern China | -5.75** | -0.10*** | -4.34** | -0.11*** |
| (Obs. = 1,503) | (2.50) | (0.03) | (1.97) | (0.04) |
| *Panel B* |  |  |  |  |
| GDP (high) | -22.70*** | -0.22*** | -13.98*** | -0.19*** |
| (Obs. = 1,053) | (5.19) | (0.05) | (3.66) | (0.05) |
| GDP (low) | -17.79*** | -0.16*** | -14.44*** | -0.16*** |
| (Obs. = 1,863) | (3.85) | (0.03) | (3.15) | (0.04) |
| per capita GDP (high) | -21.61*** | -0.21*** | -13.95*** | -0.18*** |
| (Obs. = 1,395) | (4.29) | (0.04) | (3.16) | (0.05) |
| per capita GDP (low) | -17.65*** | -0.16*** | -14.32*** | -0.16*** |
| (Obs. = 1,521) | (4.23) | (0.04) | (3.44) | (0.04) |
| Population (high) | -21.69*** | -0.22*** | -14.79*** | -0.21*** |
| (Obs. = 1,395) | (4.71) | (0.04) | (3.46) | (0.05) |
| Population (low) | -16.21*** | -0.14*** | -12.63*** | -0.12*** |
| (Obs. = 1,521) | (3.94) | (0.04) | (3.23) | (0.04) |
| Weather | Y | Y | Y | Y |
| City FE | Y | Y | Y | Y |
| Month-by-Week FE | Y | Y | Y | Y |

*Notes:* Each cell represents a separate regression using the corresponding subsample. For example, a warm region uses cities whose temperature is above the mean. Weather controls include weekly mean temperature, its square, weekly mean precipitation, and weekly mean snow depth. Standard errors are clustered at the city level and reported below the coefficients. \* significant at 10% \*\* significant at 5%. \*\*\* significant at 1%

**Table A7 (Cont.). Heterogeneity Analysis**
|  | AQI |  | PM<sub>2.5</sub> (μg/m<sup>3</sup>) |  |
| --- | --- | --- | --- | --- |
|  | levels | log | levels | log |
|  | (1) | (2) | (3) | (4) |
| *Panel C* |  |  |  |  |
| Secondary Industry Output (high) | -30.35*** | -0.27*** | -20.18*** | -0.24*** |
| (Obs. = 1,098) | (5.02) | (0.04) | (3.48) | (0.05) |
| Secondary Industry Output (low) | -11.77*** | -0.12*** | -9.46*** | -0.12*** |
| (Obs. = 1,818) | (3.77) | (0.04) | (3.08) | (0.04) |
| No. of Firms (high) | -24.74*** | -0.23*** | -16.21*** | -0.20*** |
| (Obs. = 1,188) | (4.91) | (0.04) | (3.36) | (0.04) |
| No. of Firms (low) | -13.71*** | -0.12*** | -11.05*** | -0.12*** |
| (Obs. = 1,728) | (4.02) | (0.04) | (3.38) | (0.04) |
| Transportation Activity (high) | -22.90*** | -0.24*** | -14.26*** | -0.22*** |
| (Obs. = 1,053) | (5.19) | (0.05) | (3.77) | (0.05) |
| Transportation Activity (low) | -17.17*** | -0.15*** | -13.76*** | -0.14*** |
| (Obs. = 1,863) | (3.54) | (0.03) | (2.90) | (0.03) |
| *Panel D* |  |  |  |  |
| Wastewater Emission (high) | -21.88*** | -0.24*** | -13.89*** | -0.21*** |
| (Obs. = 1,287) | (4.62) | (0.04) | (3.29) | (0.05) |
| Wastewater Emission (low) | -17.91*** | -0.14*** | -14.41*** | -0.14*** |
| (Obs. = 1,629) | (4.00) | (0.04) | (3.31) | (0.04) |
| SO<sub>2</sub> Emission (high) | -25.52*** | -0.25*** | -16.99*** | -0.22*** |
| (Obs. = 1,269) | (5.11) | (0.05) | (3.75) | (0.05) |
| SO<sub>2</sub> Emission (low) | -15.07*** | -0.13*** | -12.21*** | -0.13*** |
| (Obs. = 1,647) | (3.65) | (0.03) | (3.00) | (0.04) |
| Dust Emission (high) | -24.75*** | -0.25*** | -16.61*** | -0.22*** |
| (Obs. = 1,386) | (4.48) | (0.04) | (3.26) | (0.04) |
| Dust Emission (low) | -14.49*** | -0.12*** | -11.60*** | -0.12*** |
| (Obs. = 1,530) | (4.26) | (0.04) | (3.54) | (0.04) |
| Weather | Y | Y | Y | Y |
| City FE | Y | Y | Y | Y |
| Month-by-Week FE | Y | Y | Y | Y |

*Notes:* Each cell represents a separate regression using the corresponding subsample. For example, a warm region uses cities whose temperature is above the mean. Weather controls include weekly mean temperature, its square, weekly mean precipitation, and weekly mean snow depth. Standard errors are clustered at the city level and reported below the coefficients. \* significant at 10% \*\* significant at 5%. \*\*\* significant at 1%

**Table A8.** Summary of Related Literature on the Health Impacts of Air Pollution

| Study | Country | Period | Method | Effects |
| --- | --- | --- | --- | --- |
| *Panel A. Quasi-Experimental Studies* |  |  |  |  |
| Fan et al. (2020) | China | 2014~<br>2015 | Regression<br>Discontinuity | A 10 µg/m<sup>3</sup> increment in PM<sub>2.5</sub> is associated with a 2.2% increase in mortality rate per 100,000 using the timing of the start of winter heating. |
| He et al. (2020) | China | 2013~<br>2015 | Instrumental<br>Variable | A 10 µg/m<sup>3</sup> increment in PM<sub>2.5</sub> is associated with a 3.25% increase in mortality rate per 100,000 using the air pollution from agricultural fire. |
| Deryugina et al. (2020) | USA | 1999~<br>2013 | Instrumental<br>Variable | A 10 µg/m<sup>3</sup> increment in PM<sub>2.5</sub> is associated with a 1.8% increase in three-day mortality rate per million people aged 65+ using the wind speed. |
| *Panel B. Associational Studies* |  |  |  |  |
| Shang et al. (2013) | China | 2004~<br>2008 | Meta-Analysis | A 10-µg/m<sup>3</sup> increase in PM<sub>2.5</sub> concentrations is associated with a 0.38% increase in total mortality. |
| Zhou et al. (2015) | China | 2013 | Multi-City<br>Time-Series | A 10-µg/m<sup>3</sup> increase in two-day average PM<sub>2.5</sub> concentrations is associated with a 0.6-0.9% increase in all-cause mortality in rural China. |
| Franklin et al. (2008) | USA | 2000~<br>2005 | Hierarchical<br>Model | A 1.21% increase in all-cause mortality is associated with a 10-µg/m<sup>3</sup> increase in the previous day's PM<sub>2.5</sub> concentrations. |
| Kloog et al. (2013) | USA | 2000~<br>2008 | Time-Series | For every 10-µg/m<sup>3</sup> increase in PM<sub>2.5</sub> exposure, PM-related mortality increases by 2.8%. |
| Atkinson et al. (2014) | World | - | Meta-Analysis | A 10-µg/m<sup>3</sup> increment in PM<sub>2.5</sub> is associated with a 1.04% increase in the risk of death. The substantial regional variation is observed around the globe. |

## Footnotes

2 For example, the satellite images show dramatic declines in NO_2_ in the U.S., China, and Europe: US: https://edition.cnn.com/2020/03/23/health/us-pollution-satellite-coronavirus-scn-trnd/index.html; China: https://www.earthobservatory.nasa.gov/images/146362/airborne-nitrogen-dioxide-plummets-over-china; Europe: https://www.eea.europa.eu/highlights/air-pollution-goes-down-as. Relatedly, a news article in *Forbes* provided some rough estimates on the potential benefits caused by lockdowns in China: https://www.forbes.com/sites/jeffmcmahon/2020/03/16/coronavirus-lockdown-may-have-saved-77000-lives-in-china-just-from-pollution-reduction.

3 The explosion of the COVID-19 cases coincided with the Festival, as illustrated in Appendix Figure A3.

4 The Air Quality Index (AQI) is a comprehensive measure of air pollution in China and also widely used around the world. The index is constructed by PM_2.5_, PM_10_, SO_2_, CO, O_3_, and NO_2_ concentrations. A lower AQI means better air quality.

5 We summarize the related studies in Appendix T8.

6 For example, the morbidity cost of air pollution is found to be around two-thirds of mortality costs in China (Barwick et al., 2018).

7 For example, in Hong Kong, the number of deaths caused by influenza and the number of cases of influenza-like illness were reduced by more than 50%, as compared to previous years: https://www.ft.com/content/ad7ae6b4-5eab-11ea-b0ab-339c2307bcd4; in Japan, the influenza-like illness cases in the first week of February were only about 30% compared to the same week a year ago: https://www.japantimes.co.jp/news/2020/02/21/national/influenza-wave-drastically-wanes-japan-amid-spread-coronavirus/#.Xn9VHqgzaUl.

8 A few other studies also use quasi-experimental approaches to estimate the impacts of air pollution on mortality in China, including Chen et al. (2013), He et al. (2016), and Ebenstein et al. (2017). However, these studies are less relevant to our research context because they use data from at least a decade ago and focus on coarse measures of air pollution, such as total suspended particles (TSP), or on PM_10_.

9 We summarize the related studies in Appendix Table A8. We see that quasi-experimental studies find larger effects on mortality than associational studies.

